# Prevalence and clinical importance of titin truncating variants in adults without known congestive heart failure

**DOI:** 10.1101/19005058

**Authors:** James P. Pirruccello, Alexander Bick, Samuel Friedman, Mark Chaffin, Krishna G. Aragam, Seung Hoan Choi, Steven A. Lubitz, Carolyn Y. Ho, Kenney Ng, Anthony Philippakis, Patrick T. Ellinor, Sekar Kathiresan, Amit V. Khera

**Affiliations:** Massachusetts General Hospital Division of Cardiology, Boston, Massachusetts; Center for Genomic Medicine, Massachusetts General Hospital, Boston, Massachusetts; Program in Medical and Population Genetics at the Broad Institute, Cambridge, Massachusetts; Data Sciences Platform, Broad Institute, Cambridge, Massachusetts; Cardiovascular Research Center, Massachusetts General Hospital, Boston, Massachusetts; Cardiovascular Division, Brigham and Women’s Hospital, Boston, Massachusetts; IBM Research; Harvard Medical School, Boston, Massachusetts; Verve Therapeutics, Cambridge, Massachusetts

**Keywords:** titin, dilated cardiomyopathy, mortality

## Abstract

**Background:** Cross-sectional studies of various forms of dilated cardiomyopathy have noted a truncating mutation in the gene encoding titin (‘TTNtv’) in 7-30% of patients, but the clinical importance of identifying a TTNtv in an asymptomatic adult is largely unknown. In contrast to cross-sectional studies, prospective cohort studies allow for unbiased estimates of the disease risks associated with a genotype exposure.

**Objectives:** To determine the prevalence of cardiac imaging abnormalities and risk of incident disease among middle-aged TTNtv carriers without known congestive heart failure.

**Methods:** We analyze exome sequencing data of 45,747 participants of the UK Biobank without known congestive heart failure to identify TTNtv carriers. Among 10,552 with cardiac magnetic resonance imaging (MRI), we determine the relationship between TTNtv carrier status and left ventricular ejection fraction. In this prospective cohort, we quantify the absolute and relative risks of incident disease in TTNtv carriers versus noncarriers.

**Results:** Among 45,747 middle-aged participants without known congestive heart failure, 196 (0.43%) harbored a TTNtv. The average ejection fraction was 61% in TTNtv carriers versus 65% in noncarriers (P = 1.8 × 10^−8^), with a 9.3-fold increase (95% CI 3.9 – 22.2) in odds of subnormal ejection fraction (P = 5.7 × 10^−5^). Over a median follow-up of 6.9 years, a composite endpoint of incident dilated cardiomyopathy, congestive heart failure, or all-cause mortality was observed in 6.6% of TTNtv carriers versus 2.9% of non-carriers (adjusted hazard ratio 2.5; 95% CI 1.4 – 4.3; p = 1.1 × 10^−3^).

**Conclusions:** Approximately 1 in 230 middle-aged adults without known congestive heart failure harbored a TTNtv. These carriers had a substantially increased relative risk—but modest absolute risk—of having a subnormal ejection fraction or manifesting clinical disease during prospective follow-up.

**Condensed Abstract:** Cross-sectional studies of dilated cardiomyopathy have noted a truncating mutation in the gene encoding titin (‘TTNtv’) in up to 30% of patients—but the clinical importance of TTNtv in asymptomatic adults is largely unknown. Here, we observe a TTNtv in 0.43% of 45,747 middle-aged adults. Average ejection fraction was 61% in TTNtv carriers versus 65% in non-carriers (p<0.001). Over a median follow-up of 7 years, incident congestive heart failure or mortality was observed in 6.6% of TTTtv carriers versus 2.9% of non-carriers (hazard ratio 2.5; p = 0.001).

## Text

### Introduction

Truncating variants in the gene encoding titin (TTNtv) are the most commonly identified mutations in patients presenting with dilated cardiomyopathy (DCM)(1–4). In principle, gene sequencing to identify asymptomatic individuals who harbor a TTNtv could enable early diagnosis or preventive therapy. In practice, the clinical importance of identifying a TTNtv in an asymptomatic individual with respect to relative and absolute risks of incident disease is largely unknown.

A seminal paper in 2012 documented a TTNtv in ∼20% of patients with DCM, a finding subsequently replicated across a range of important forms of DCM—peripartum (15%), alcohol-induced (14%), and chemotherapy related (8%)(1–4). As compared to controls, these studies suggested a 5-10 fold increased odds of developing disease among TTNtv carriers. Among individuals without clinically manifest disease, small prior studies have suggested a modestly reduced ejection fraction among TTNtv carriers(5, 6). More recently, the increased prevalence of TTNtv in patients with DCM was replicated in cross-sectional analyses from biobanks of two large U.S. health systems(7).

Gene sequencing to identify individuals with important genetic variants at scale in the population has become increasingly practical as the costs of sequencing have declined rapidly. At least one U.S. healthcare system has already started returning TTNtv carriers status to patients within routine clinical practice (Dewey et al., 2016), and a similar commitment has been embraced as a core tenet of the All of Us Research Program, a National Institutes of Health sponsored program that plans to enroll over one million participants in coming years(8).

Here, we determine the clinical importance of identifying a TTNtv among middle-aged adults of the UK Biobank without known congestive heart failure. We first determined the relationship between TTNtv carrier status and subclinical cardiac imaging abnormalities. Second, we characterize the relative and absolute risks of incident congestive heart failure or all-cause mortality in prospective follow-up.

## Methods

### Study participants

The UK Biobank is a richly phenotyped, prospective cohort of over 500,000 individuals (9). Participants were enrolled between 2006-2010, and were aged 40-69 at the time of enrollment. Analysis of cardiac MRI phenotypes was performed in all individuals with both genetic and imaging data, after exclusion of participants with a cardiomyopathy or coronary artery disease diagnosis prior to the date of imaging. Incident event analyses were performed in all available participants, after exclusion of those with a cardiomyopathy or coronary artery disease diagnosis prior to enrollment.

Analysis of the UK Biobank data was approved by the Partners HealthCare institutional review board (protocol 2013P001840). Work was performed under UK Biobank application #7089.

### Exome sequencing and *TTN* truncating variant classification

Whole exome sequencing was performed in 49,997 participants as previously described(10, 11). 53 participants without genotyping array data were removed, leaving 49,944 exomes for downstream analysis.

TTNtv variants were classified using the LOFTEE plug-in of the Ensembl Variant Effect Predictors(12, 13). This algorithm has been previously shown to reliably classify inactivating variants: those that lead to a premature stop codon (nonsense), disrupt a canonical exon splicing consensus sequence, or cause an inactivating frameshift. Unless otherwise noted, all subsequent analyses were conducted using TTNtv in exons presents in cardiac-specific transcripts with percent spliced in (PSI) in cardiac transcripts greater than 90% as previously recommended(5).

### Phenotype definitions

Disease phenotypes in the UK Biobank were defined using a combination of self-reported data (confirmed by a healthcare professional), and hospital admission diagnosis codes, and death registry data. We defined DCM to be International Statistical Classification of Diseases (ICD-10) code I42.0, in the absence of a history of coronary artery disease (CAD), coronary artery bypass grafting, or percutaneous coronary intervention as previously reported(14). Additional details are provided in **Supplemental Table 1**.

**Table 1:**
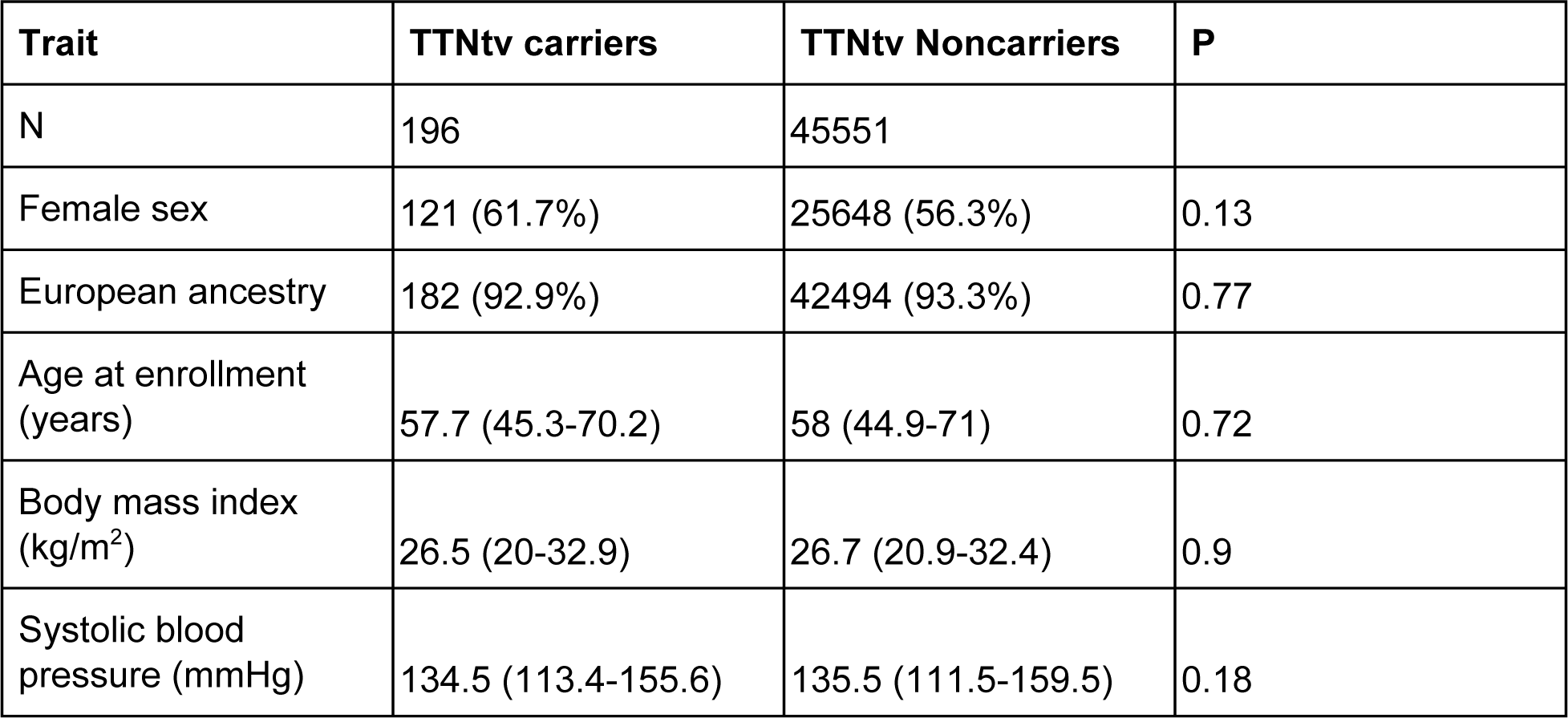
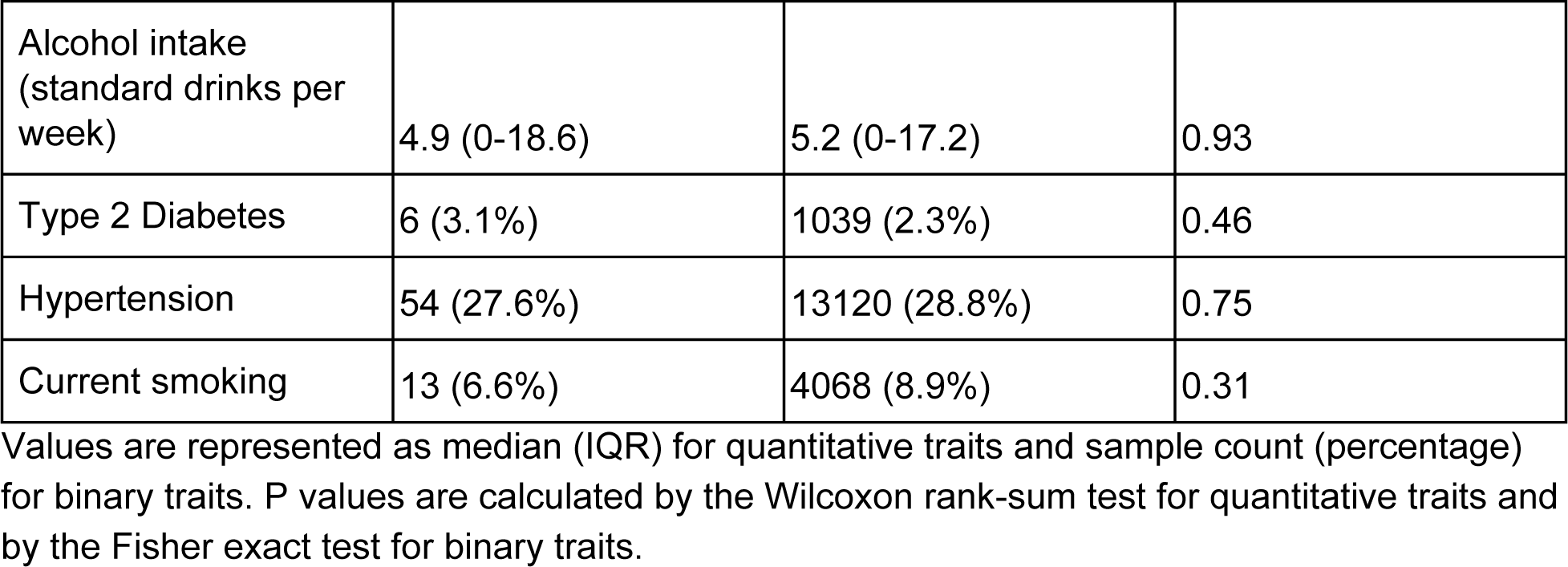
Baseline characteristics.

### Cardiac magnetic resonance imaging measurements

Among the 45,747 individuals included in the main analysis, 10,784 had cardiac MRI data available. These individuals were invited for imaging by the UK Biobank based on proximity to imaging centers, without consideration of disease status. Cardiac MRI was performed with 1.5 Tesla machines (MAGNETOM Aera, Syngo Platform InlineVF version D13A, Siemens Healthcare) with electrocardiographic gating for cardiac synchronization (15). Cardiac assessment was performed from the combination of several cine series using balanced steady-state free precession acquisitions (16). Automated readings were produced for left ventricular end diastolic volume (LVEDV), left ventricular end systolic volume (LVESV), and left ventricular ejection fraction (LVEF). LVEDV and LVESV were corrected with previously established linear equations tailored to a UK population on the same imaging platform (17).

A cardiologist (JPP) reviewed the automated tracings from imaging studies belonging to individuals with outlier measurements, defined as samples outside of 1.5 interquartile ranges below the first or above the third quartile of LVEDV or LVESV (18). 232 automated interpretations were deemed to be anatomically mistraced, leaving 10,552 individuals with cardiac MRI data for analysis. In the remaining samples, LVEF below 54% for women or 52% for men was considered to be subnormal (19).

### Statistical analysis

To assess the association between TTNtv and prevalent disease, we applied a Firth bias-corrected logistic regression model adjusted for sex and genetic ancestry as quantified by the first five principal components, derived centrally by the UK Biobank as previously reported(9, 20). For assessing the association between TTNtv and subnormal LVEF, we applied Firth bias corrected logistic regression adjusted for the cubic basis spline of age at enrollment (in order to adjust for non-linear effects of age(21)), age at the time of MRI, sex, and the first five principal components of ancestry.

Incident disease analyses were conducted using Cox proportional hazard models adjusted for the cubic basis spline of age at the start of follow-up time, sex, and the first five principal components of ancestry. Start of follow-up time was considered to be the date of enrollment or—to prevent confounding introduced by ‘immortal time bias’(22)—the date of cardiac MRI for those who underwent imaging. Additional details are available in the **Supplementary Methods**.

Statistical analyses were conducted using R version 3.4.4 (R Foundation for Statistical Computing, Vienna, Austria). Unadjusted comparison of baseline characteristics between TTNtv carriers and noncarriers was performed with the Wilcoxon rank sum test for quantitative traits and with the Fisher exact test for binary traits. Two-sided p-values were used unless otherwise specified.

## Results

### TTNtv variant identification

49,997 participants had exome sequencing data, of whom 53 were excluded due to unavailable genotyping array data needed for genetic ancestry assessment. In the remaining 49,944 individuals (before applying disease-based sample exclusion criteria), 308 distinct TTNtv variants were identified across 669 carriers. Of these, 178 TTNtv variants were found in exons highly expressed in the heart (percent spliced in > 90%, “high-PSI”), including 64 stop-gain variants, 87 frameshifts, and 27 splice-site disrupting variants. These high-PSI variants were present in a total of 227 individuals (0.45%) (**Figure 1**).

**Figure 1:**
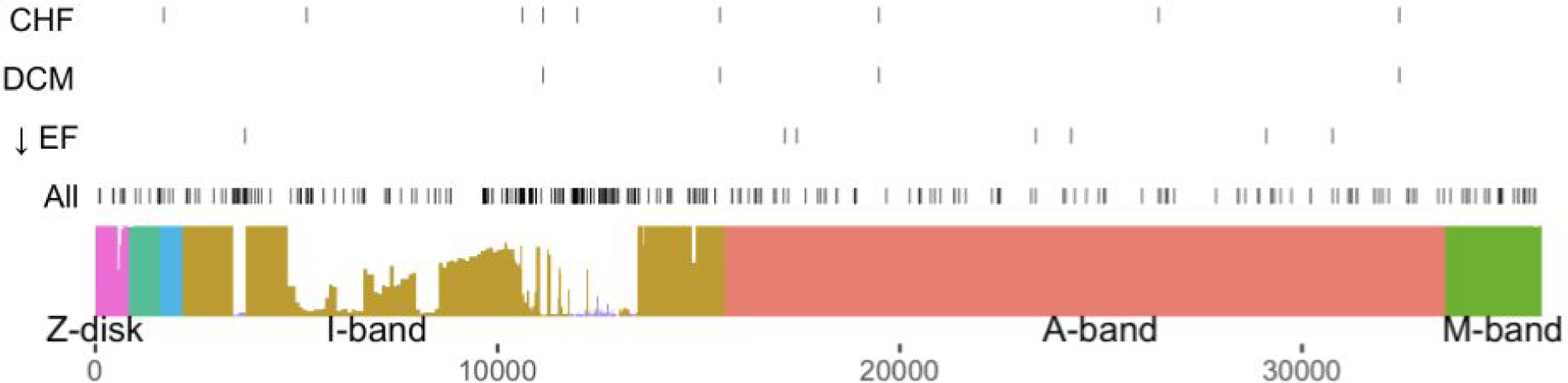
*TTN* truncating variants. Domains of TTN, including the Z-disk, the I-band, the A-band, and the M-band, are colored and labeled. The distance along the TTN transcript is demarcated on the x-axis. Each exon is scaled on the y-axis based on its PSI value, reflecting the percentage of *TTN* transcripts found in the heart that incorporate the exon (5). Each hash mark represents one individual with a TTNtv, based on the location of that truncation; the top row of hash marks represents individuals who developed incident congestive heart failure; the second, those who developed incident dilated cardiomyopathy; the third, those with a reduced LVEF; and the fourth, all other individuals with a TTNtv. Individuals with TTNtv in any exon, including those with PSI < 90%, are represented.

### TTNtv were associated with prevalent CHF and DCM

To confirm prior case-control observations, we first assessed the relationship between high-PSI TTNtv and prevalent disease in 227 high-PSI TTNtv carriers and 49,717 noncarriers. Four TTNtv carriers had DCM at baseline compared to 22 noncarriers (adjusted odds ratio [OR] 51.5, 95% CI 19.0-139.6, P = 4.1 × 10^−7^). 10 TTNtv carriers had CHF, compared to 302 noncarriers (OR 8.0, 95% CI 4.2-15.0, P = 7.7 × 10^−7^). As expected, the presence of a TTNtv in an exon not highly expressed in the heart (PSI < 90%) was not significantly associated with prevalent disease among 421 carriers (P = 0.58 for DCM; P = 0.61 for CHF).

### Participant baseline characteristics for incident disease analysis

For subsequent analyses, we excluded participants with CHF, DCM, or CAD at baseline (**Figure 2**), after which 45,747 participants remained. Of these, 196 (0.43%) had a high PSI TTNtv. Baseline characteristics were similar between participants with and without TTNtv (**Table 1** and **Supplementary Table 2**).

**Table 2:** Cardiac MRI characteristics.

| Trait | TTNtv carriers | TTNtv Noncarriers | P |
| --- | --- | --- | --- |
| N | 47 | 10,505 |  |
| Women | 29 (61.7%) | 5571 (53%) |  |
| Age at time of MRI (years) | 61.3 (53.8-68.8) | 62.3 (54.8-69.8) |  |
| LVEF (%) | 61.1% (53.7%-68.5%) | 64.9% (59.4%-70.5%) | $1.8 \times 10^{-8}$ |
| LVESV (mL) | 56.0 (33.1-79.0) | 49.2 (33.5-64.8) | $6.6 \times 10^{-6}$ |
| LVEDV (mL) | 140.2 (108.0-172.3) | 138.0 (109.8-166.1) | 0.11 |

Values are represented as mean ( ± SD) for quantitative traits and sample count (percentage) for binary traits. P represents the TTNtv P-value from a linear model adjusting for the cubic basis spline of age, sex, and the first five principal components of ancestry.

**Figure 2:**
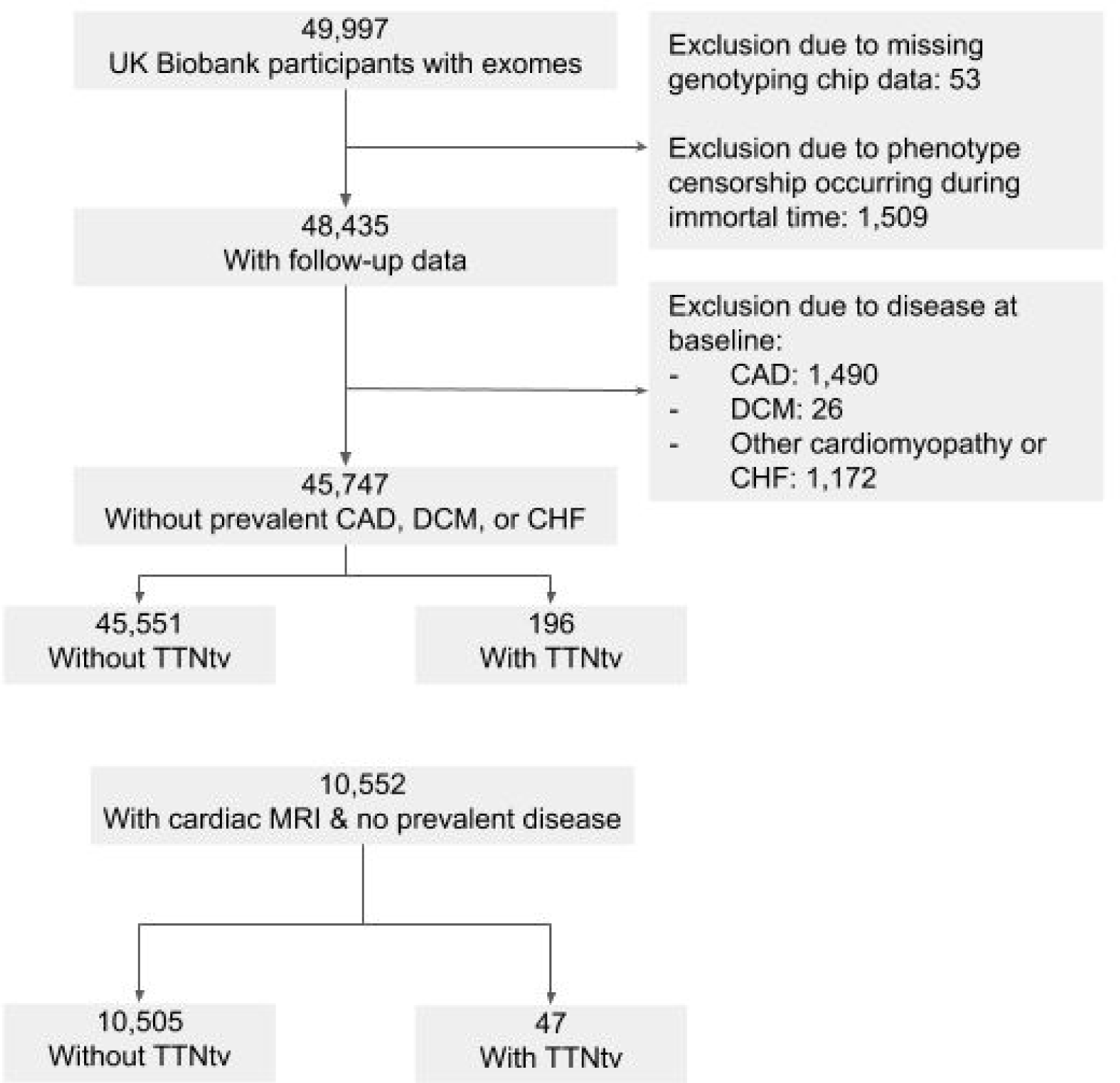
Flow diagram of samples included in the study. TTNtv: Titin truncating variants in exons spliced into greater than 90% of transcripts in the heart. CAD: Coronary artery disease. DCM: Dilated cardiomyopathy. CHF: Congestive heart failure. MRI: Magnetic resonance imaging.

### TTNtv were linked to subclinical changes in cardiac structure and function

We next examined the relationship between TTNtv and subclinical cardiac MRI abnormalities in 10,552 without known CAD, DCM, or CHF at time of imaging. Of these participants, 47 (0.45%) carried a TTNtv. Importantly, the UK Biobank invited individuals to undergo imaging without consideration of disease status, thus minimizing potential biases introduced by imaging ordered as part of routine clinical care.

Mean LVEF was 61.1% in TTNtv carriers versus 64.9% in noncarriers (adjusted difference −4.3%; 95% CI −2.8% – −5.8%; P = 1.8 × 10^−8^). Significantly more TTNtv carriers had a subnormal LVEF, defined as being below 52% for men or 54% for women (19), compared to noncarriers (13% vs 2.1%; **Figure 3**). In an adjusted logistic regression model, the presence of a TTNtv yielded an OR of 9.3 (CI 3.9-22.2; P = 5.7 × 10^−5^) for a subnormal ejection fraction.

**Figure 3:**
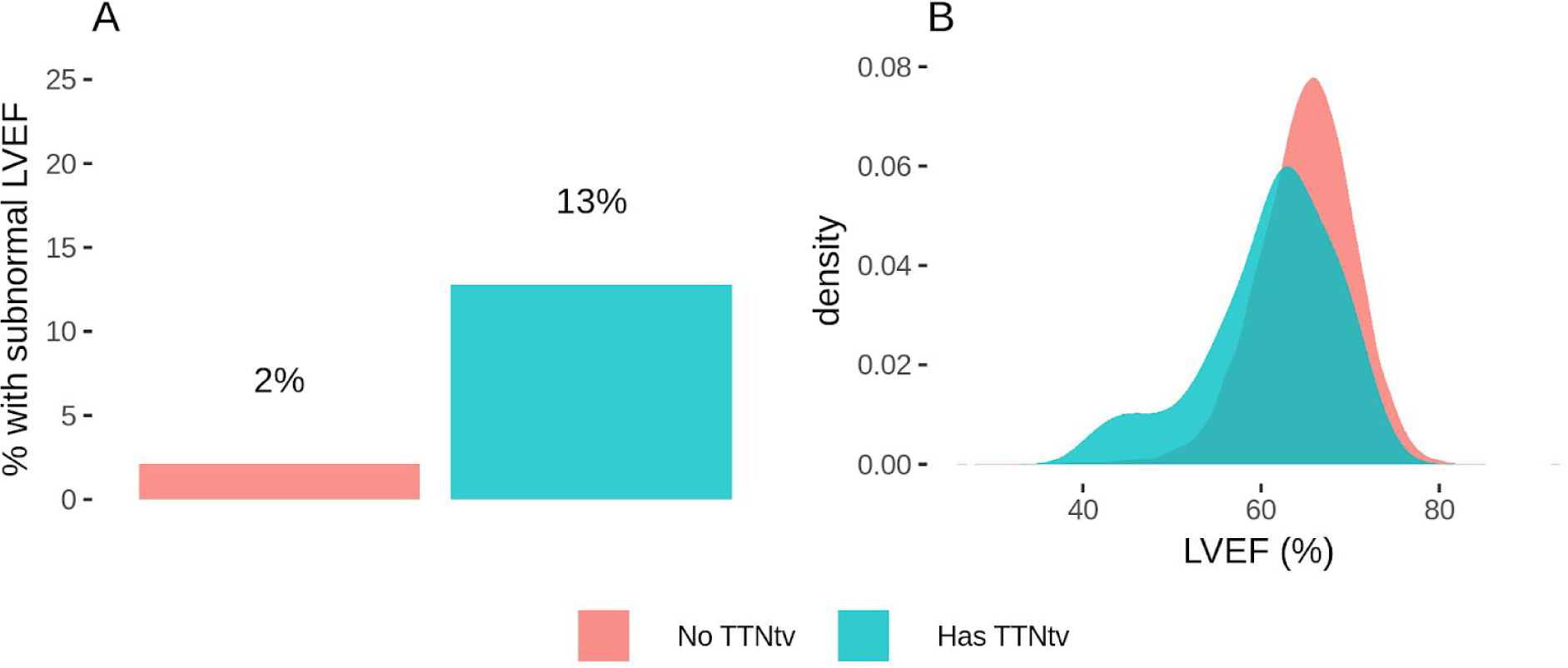
Impact of TTNtv carrier status on left ventricular ejection fraction. In Panel **A**, the proportion of the population with a subnormal LVEF is shown, stratified into TTNtv carriers and noncarriers. Men with LVEF below 0.52 and women with LVEF below 0.54 were considered to have a subnormal LVEF. The presence of a TTNtv was associated with increased odds of having a subnormal LVEF (OR 9.3; P = 5.7 × 10^−5^) in a logistic model adjusted for the cubic basis spline of age, sex, and the first five principal components of ancestry. In Panel **B**, the distribution of LVEF is shown in TTNtv carriers versus noncarriers. Mean LVEF was 61.1% in TTNtv carriers versus 64.9% in noncarriers, corresponding to a −4.3% adjusted difference (P = 1.8 × 10^−8^) in a linear model accounting for the cubic basis spline of age, sex, and the first five principal components of ancestry.

Beyond LVEF, TTNtv carriers had no significant difference in LV end-diastolic volume but did have a significantly increased LV end-systolic volume (adjusted difference 8.7mL; 95% CI 4.9-12.4; P = 6.6 × 10^−6^), consistent with impaired contractility among TTNtv carriers.

### TTNtv were associated with incident morbidity and mortality

Among 45,747 individuals without clinically diagnosed CHF or coronary artery disease at baseline, we next determined relative and absolute risk of incident DCM, CHF, or all-cause mortality over a median follow-up of 6.9 years (25th-75th% 6.3-7.1 years). The composite endpoint of a physician diagnosis code for DCM, CHF, or mortality occurred in 13 of 196 (6.6%) TTNtv carriers and 1,329 of 45,551 (2.9%) of noncarriers—corresponding to an adjusted hazard ratio (HR) of 2.5 (95% CI 1.4-4.3; P = 1.1 × 10^−3^). The increased HR for the composite outcome in TTNtv carriers was not attenuated by adjusting for additional clinical risk factors at baseline, including alcohol consumption, smoking status, hypertension, diabetes, and BMI (adjusted HR 2.5, 95% CI 1.5-4.4, P = 8.9 × 10^−4^).

With respect to individual components of the composite endpoint, 3 out of 196 TTNtv carriers developed incident DCM, compared to 23 of 45,551 noncarriers (adjusted HR 31.7, 95% CI 9.5-106.5; P=2.2 × 10^−8^). 5 of 196 TTNtv carriers developed CHF, compared to 277 of 45,551 noncarriers (adjusted HR 4.1; 95% CI 1.7-9.8; P=1.9 × 10^−3^). 9 of 196 carriers died, compared to 1,115 of 45,551 noncarriers (adjusted HR 1.9; 95% CI 1.0-3.7; P=0.052; **Supplementary Figure 1**). The increased risk of mortality for TTNtv carriers was not attenuated by adjusting for additional clinical risk factors at baseline, including alcohol consumption, smoking status, hypertension, diabetes, and BMI (adjusted HR 1.9, 95% CI 1.0-3.7, P=0.047).

Similar patterns were observed in an analysis that quantified 5-year risk of each of the endpoints (**Figure 4**).

**Figure 4:**
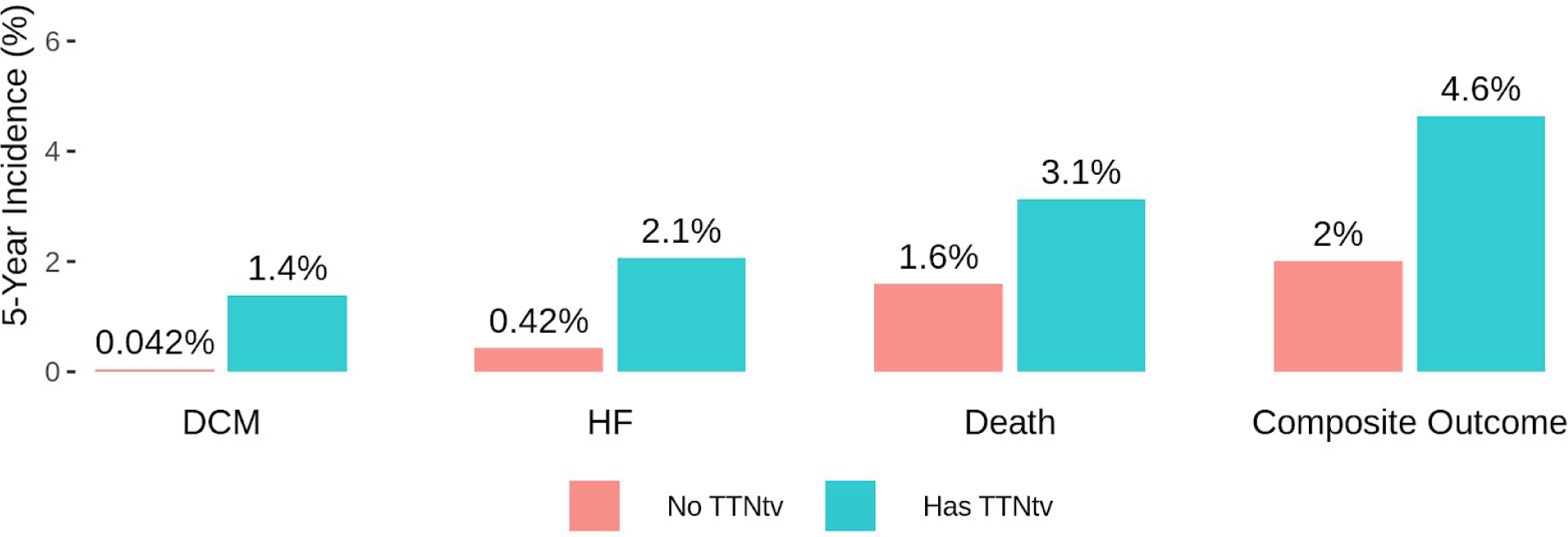
Relationship between TTNtv and 5-year incidence of dilated cardiomyopathy, heart failure, and mortality. The percentages reflect the incident event curve modeled at 5 years for the development of DCM, CHF, death, or the composite outcome (DCM, CHF, or death), stratified according to TTNtv carrier status.

## Discussion

A primary goal of genomic medicine is to identify asymptomatic individuals who are nonetheless high-risk based on genetic variants, with the aim of facilitating targeted screening or prevention efforts. This paradigm is a significant departure from the traditional approach, where gene sequencing has been largely restricted to those already afflicted with disease and their family members. Here, we observe a TTNtv in 1 in ∼230 middle-aged adult participants without known cardiovascular disease. On average, TTNtv carriers had subtly lower LVEF (61% vs 65%, p=1.8 × 10^−8^) and were 9-fold more likely to have subnormal LVEF. During a median follow-up period of 6.9 years, TTNtv carriers had a high relative risk of developing DCM, CHF, or death (HR 2.5, 95% CI 1.7-3.6), but a modest absolute risk (6.6% among carriers vs 2.9% among noncarriers).

The results of these analyses support three key conclusions. First, beyond confirming a known association of TTNtv variants with prevalent cardiomyopathy and CHF (2, 3, 5, 7, 23–28), we provide new evidence for a substantially increased risk of incident DCM, CHF, and all-cause mortality. This prospective assessment is particularly important in helping to overcome concerns about ascertainment biases that may be present in prior seminal case-control studies.

Second, despite a substantially increased relative risk noted in TTNtv carriers, the absolute risk of an abnormal ejection fraction and incident CHF remains modest. For example, 87% of the TTNtv carriers who underwent cardiac imaging in our study had a normal ejection fraction and 97% remained free of a new clinical diagnosis of CHF after a median follow-up of 6.9 years. These results are consistent with TTNtv serving as an important risk factor for cardiomyopathy, rather than mapping with disease in a 1:1 deterministic fashion. Work from prior studies suggests that superimposed clinical or environmental stressors—pregnancy, alcohol, or chemotherapy—may unmask overt cardiomyopathy among those genetically predisposed by a TTNtv(1–4). Additional efforts are needed to better understand the genetic and nongenetic determinants governing which TTNtv carriers are most likely to manifest clinical disease.

Third, *TTN* may warrant inclusion in the American College of Medical Genetics and Genomics list of genes in which pathogenic mutations are both important and actionable (29). Although the management of asymptomatic TTNtv carriers lacks a strong evidence base, potential clinical considerations include serial imaging to detect subclinical structural abnormalities, avoidance of excess alcohol, intensive blood pressure control, increased surveillance or dose-reduction of cardiotoxic chemotherapeutic regimens, and ‘cascade screening’ to assess carrier status in first-degree relatives (30). At least one U.S. health system has already started disclosing TTNtv variant status to patients and their health care providers (31); this experience is likely to provide further insights into the clinical utility, as well as potential adverse psychosocial or cost implications of this approach.

These results are best interpreted in the context of four key limitations. First, the detection of prevalent and incident CHF-related outcomes was based on verbal interviews with healthcare staff from the UK Biobank and hospital admission diagnosis codes—as such, there may be inaccuracies in clinical event adjudication. Second, UK Biobank participants were recruited at age 40-69 years, raising the possibility of survivorship or selection biases, which limits the generalizability of our results to younger or elderly patients. Third, cardiac MRI data were available in a subset of the participants and at one point in time for each participant; analyses of serial cardiac imaging studies would be informative to model trajectories in cardiac function over time. Fourth, >90% of the participants in our study were of European ancestry, limiting the ability to assess for effect heterogeneity across other ancestral groups, as was suggested in a recent analysis of black patients of U.S. health care systems (7).

## Conclusions

In conclusion, we identified TTNtv variants in 0.43% of individuals without known congestive heart failure. These individuals had subtle, subnormal reductions in LV systolic function and were at substantially increased relative risk—but modest absolute risk—of developing incident dilated cardiomyopathy, congestive heart failure, or all-cause mortality.

## Data Availability

Data is available to researchers from the UK Biobank following their standard access request procedures.

## Abbreviations

MRI: magnetic resonance imaging
TTNtv: titin truncating variants
DCM: dilated cardiomyopathy
CHF: congestive heart failure
CAD: coronary artery disease
LVEF: left ventricular ejection fraction
LVEDV: left ventricular end diastolic volume
LVESV: left ventricular end systolic volume
ICD: International Statistical Classification of Diseases.

## References

1. Garcia-Pavia P, Kim Y, Alejandra Restrepo-Cordoba M, et al. Genetic Variants Associated with Cancer Therapy-Induced Cardiomyopathy. Circulation 2019.

2. Herman DS, Lam L, Taylor MRG, et al. Truncations of Titin Causing Dilated Cardiomyopathy. N. Engl. J. Med. 2012;366:619–628.

3. Ware JS, Li J, Mazaika E, et al. Shared Genetic Predisposition in Peripartum and Dilated Cardiomyopathies. N. Engl. J. Med. 2016;374:233–241.

4. Ware JS, Amor-Salamanca A, Tayal U, et al. Genetic Etiology for Alcohol-Induced Cardiac Toxicity. J. Am. Coll. Cardiol. 2018;71:2293–2302.

5. Roberts AM, Ware JS, Herman DS, et al. Integrated allelic, transcriptional, and phenomic dissection of the cardiac effects of titin truncations in health and disease. Sci. Transl. Med. 2015;7:270ra6.

6. Schafer S, de Marvao A, Adami E, et al. Titin-truncating variants affect heart function in disease cohorts and the general population. Nat. Genet. 2017;49:46–53.

7. Haggerty Christopher M., Damrauer Scott M., Levin Michael G., et al. Genomics-First Evaluation of Heart Disease Associated With Titin-Truncating Variants. Circulation 2019;140. Available at: https://www.ahajournals.org/doi/abs/10.1161/CIRCULATIONAHA.119.039573. Accessed June 20, 2019.

8. Denny JC, Rutter J, Goldstein DB, et al. The “All of Us” Research Program. N. Engl. J. Med. 2019;381:668–676.

9. Bycroft C, Freeman C, Petkova D, et al. The UK Biobank resource with deep phenotyping and genomic data. Nature 2018;562:203.

10. Hout CVV, Tachmazidou I, Backman JD, et al. Whole exome sequencing and characterization of coding variation in 49,960 individuals in the UK Biobank. bioRxiv 2019:572347.

11. Regier AA, Farjoun Y, Larson DE, et al. Functional equivalence of genome sequencing analysis pipelines enables harmonized variant calling across human genetics projects. Nat. Commun. 2018;9:4038.

12. Karczewski KJ, Francioli LC, Tiao G, et al. Variation across 141,456 human exomes and genomes reveals the spectrum of loss-of-function intolerance across human protein-coding genes. bioRxiv 2019:531210.

13. McLaren W, Gil L, Hunt SE, et al. The Ensembl Variant Effect Predictor. Genome Biol. 2016;17:122.

14. Aragam Krishna G, Chaffin Mark, Levinson Rebecca T, et al. Phenotypic Refinement of Heart Failure in a National Biobank Facilitates Genetic Discovery. Circulation 2018;0. Available at: https://www.ahajournals.org/doi/10.1161/CIRCULATIONAHA.118.035774. Accessed December 9, 2018.

15. Petersen SE, Matthews PM, Francis JM, et al. UK Biobank’s cardiovascular magnetic resonance protocol. J. Cardiovasc. Magn. Reson. 2016;18. Available at: https://www.ncbi.nlm.nih.gov/pmc/articles/PMC4736703/. Accessed November 18, 2018.

16. Petersen SE, Aung N, Sanghvi MM, et al. Reference ranges for cardiac structure and function using cardiovascular magnetic resonance (CMR) in Caucasians from the UK Biobank population cohort. J. Cardiovasc. Magn. Reson. 2017;19:18.

17. Sanghvi MM, Feuchter P, Zemrak F, et al. Automatic left ventricular analysis with Inline VF performs well compared to manual analysis: results from Barts Cardiovascular Registry. J. Cardiovasc. Magn. Reson. 2016;18. Available at: https://www.ncbi.nlm.nih.gov/pmc/articles/PMC5032600/. Accessed June 1, 2019.

18. Tukey JW. Exploratory data analysis. Reading, Mass.: Addison-Wesley Pub. Co.; 1977.

19. Lang RM, Badano LP, Mor-Avi V, et al. Recommendations for Cardiac Chamber Quantification by Echocardiography in Adults: An Update from the American Society of Echocardiography and the European Association of Cardiovascular Imaging. J. Am. Soc. Echocardiogr. 2015;28:1-39.e14.

20. Firth D. Bias reduction of maximum likelihood estimates. Biometrika 1993;80:27–38.

21. Jr FEH. Regression Modeling Strategies: With Applications to Linear Models, Logistic and Ordinal Regression, and Survival Analysis. Springer; 2015.

22. Secemsky EA, Yeh RW. Complete vs Incomplete Revascularization During Percutaneous Coronary Intervention and Improved Survival—The Key Is Immortality. JAMA Cardiol. 2018;3:443–444.

23. Akinrinade O, Ollila L, Vattulainen S, et al. Genetics and genotype-phenotype correlations in Finnish patients with dilated cardiomyopathy. Eur. Heart J. 2015;36:2327–2337.

24. Franaszczyk M, Chmielewski P, Truszkowska G, et al. Titin Truncating Variants in Dilated Cardiomyopathy – Prevalence and Genotype-Phenotype Correlations. PLOS ONE 2017;12:e0169007.

25. Gerull B, Gramlich M, Atherton J, et al. Mutations of TTN, encoding the giant muscle filament titin, cause familial dilated cardiomyopathy. Nat. Genet. 2002;30:201–204.

26. Norton N, Li D, Rampersaud E, et al. Exome sequencing and genome-wide linkage analysis in 17 families illustrate the complex contribution of TTN truncating variants to dilated cardiomyopathy. Circ. Cardiovasc. Genet. 2013;6:144–153.

27. van Spaendonck-Zwarts KY, Posafalvi A, van den Berg MP, et al. Titin gene mutations are common in families with both peripartum cardiomyopathy and dilated cardiomyopathy. Eur. Heart J. 2014;35:2165–2173.

28. Tayal U, Newsome S, Buchan R, et al. Phenotype and Clinical Outcomes of Titin Cardiomyopathy. J. Am. Coll. Cardiol. 2017;70:2264–2274.

29. Kalia SS, Adelman K, Bale SJ, et al. Recommendations for reporting of secondary findings in clinical exome and genome sequencing, 2016 update (ACMG SF v2.0): a policy statement of the American College of Medical Genetics and Genomics. Genet. Med. Off. J. Am. Coll. Med. Genet. 2017;19:249–255.

30. Hershberger RE, Givertz MM, Ho CY, et al. Genetic Evaluation of Cardiomyopathy—A Heart Failure Society of America Practice Guideline. J. Card. Fail. 2018;24:281–302.

31. Dewey FE, Murray MF, Overton JD, et al. Distribution and clinical impact of functional variants in 50,726 whole-exome sequences from the DiscovEHR study. Science 2016;354:aaf6814.

